# Long-Term Outcome after Convergent Procedure for Atrial Fibrillation

**DOI:** 10.1101/2023.11.20.23298797

**Authors:** Borut Gersak, Veronika Podlogar, Tine Prolic Kalinsek, Matevz Jan

## Abstract

**Background:** The aim of this single-center retrospective study is to evaluate the long-term outcome after convergent procedure (CP) for patients with paroxysmal atrial fibrillation (AF), persistent AF and long-standing persistent AF.

**Methods and results:** We analyzed outcomes of patients that underwent CP from January 2009 until July 2020. 119 patients with paroxysmal AF (23.5%), persistent AF (5.9%) or long-standing persistent AF (70.6%) that attended long-term follow up were included. The outcome was assessed at 1-year after CP and at long-term follow up. At 1-year follow up rhythm and daily AF burden were assessed for patients with implantable loop recorder (61.2%). For others rhythm was assessed by clinical presentation and 12-lead ECG recording. At long-term follow up patients having sinus rhythm or unclear history of AF were monitored with 7-day ECG Holter and AF burden was determined. Long-term success was defined as freedom from AF/atrial flutter (AFL) with sinus rhythm on 12-lead ECG recording and AF/AFL burden <1% on 7-day Holter ECG. Repeat catheter ablations (RFA) prior to long-term follow up were documented.

At 1-year follow up 91.4% of patients had sinus rhythm and 76.1% of patients had AF/AFL burden <1%. At long-term follow up (8.3 ± 2.8 years) 65.5% of patients had sinus rhythm and 53.8% patients had AF/AFL burden <1% on 7-day holter ECG. Additional RFAs were performed in 32.8% of patients who had AF or AFL burden <1%. At long-term follow up age, body mass index and left atrial volume index were associated with increased risk of AF recurrence.

**Conclusions:** CP resulted in high long-term probability of sinus rhythm maintenance. During long-term follow-up additional RFAs were required to maintain sinus rhythm in a substantial number of patients.

## Introduction

Atrial fibrillation (AF) is a worldwide epidemic that results in significant burden on public health and health care system (1,2). The incidence and prevalence of AF is consistently increasing throughout the years (2,3). Patients with AF have increased risk of heart failure, myocardial infarction, stroke, dementia, and mortality (4,5). Higher AF burden decreases health-related quality of life (6–8).

Several treatments for AF have been developed with catheter ablation for pulmonary vein isolation (PVI) being a Class IA indication for paroxysmal AF treatment (9). However, more successful in improving the outcomes of AF ablation than medical therapy, the success rate of catheter ablation for persistent and longstanding persistent AF is to this date still far from being adequate (10,11).

Convergent procedure (CP) is a multidisciplinary solution for treatment of AF that involves epicardial ablation of the left atrium with minimally invasive surgical approach and endocardial catheter ablation. It was demonstrated to be superior in treating persistent AF compared to endocardial-only ablation strategy in the CONVERGE trial (12,13). Additionally, it was shown to be superior in improving the outcomes in patients with paroxysmal AF (14,15).

However, few studies were published exploring the long-term outcomes of the CP (12,16–18). The objective of our retrospective analysis is to evaluate the very long-term outcome for patients with paroxysmal, persistent, and longstanding persistent AF that underwent CP.

## Methods

### Patients

Our single-center retrospective analysis reports long-term outcome for patients that underwent CP from January 2009 until July 2020. During this period 170 adult patients with symptomatic paroxysmal, persistent, or longstanding persistent AF received CP at the University Medical Centre Ljubljana. The CP protocol was reviewed and approved by the National Medical Ethics Committee.

### Convergent Procedure

CP was performed either at the same hospitalization as a single-setting procedure or as sequential epicardial first, endocardial later procedure on two different hospitalizations. When single setting was utilized, percutaneous endocardial ablation was performed immediately after the epicardial ablation was completed.

The procedure was performed under general anesthesia in hybrid operating room equipped with fluoroscopy and electrophysiology mapping system. After February 2010 temperature probe was placed in the esophagus and temperature was monitored during both epicardial and endocardial procedure, as well as ice cold saline was used intrapericardially to cool the ablation area further.

### Epicardial Ablation

The access to pericardial space was obtained through laparoscopic transabdominal transdiaphragmatic approach or subxiphoid approach. Later, a subxiphoid, transthoracic approach was utilized.

In transdiaphragmatic approach access to posterior wall of left atrium was obtained under CO2 insufflation into the abdominal cavity using laparoscopic instruments inserted through three abdominal trocars as described previously (19). Pericardial window was created through the central tendon of the diaphragm and pericardium. The Subtle™ cannula (Atricure, Mason, OH, USA/nContact, Morrisville, NC, USA) was inserted into the oblique sinus, providing a direct view of the posterior left atrium through endoscope. The epicardial radiofrequency (RF) ablation device Numeris^®^ or Episense™ (both Atricure, Mason, OH, USA/nContact, Morrisville, NC, USA) was introduced through the cannula, adjacent to the endoscope. Under direct endoscopic visualization interconnected epicardial lesions were created, as shown in Figure 1. During epicardial ablation ice cold saline was injected into the pericardial sac since February 2010 to minimize heating of adjacent structures and periprocedural complications.

**Fig. 1.**
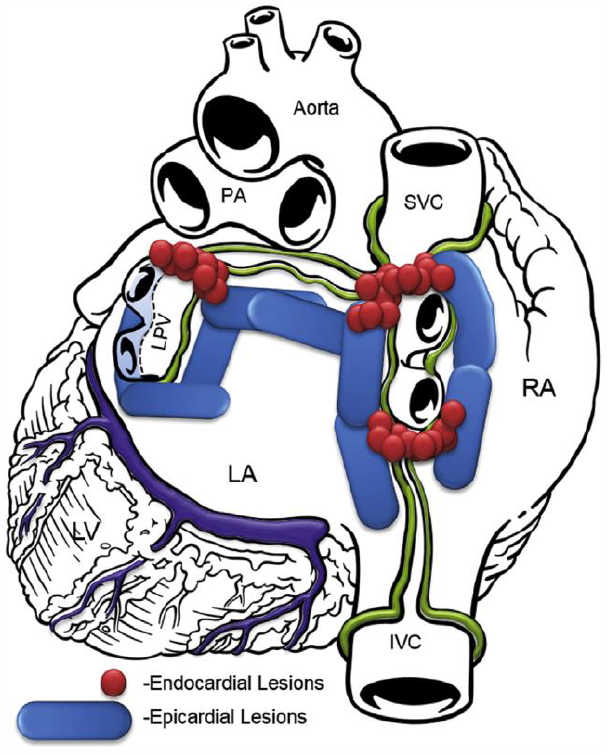

In subxiphoid approach incision was made directly under the sternum and the pericardial space was entered just above the diaphragm. Procedure was performed similarly as in transabdominal approach. Since 2015 the lesion set was focused more on ablating the whole posterior left atrium as shown in Figure 2.

**Fig. 2.**
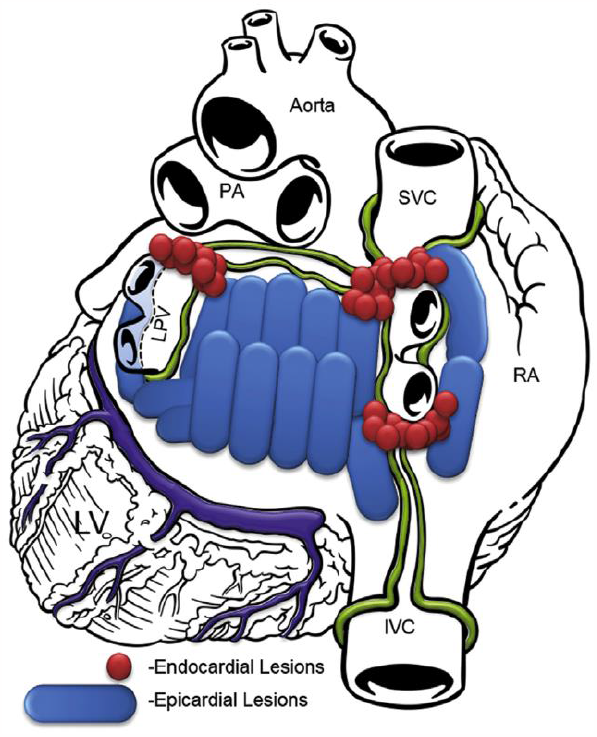

After epicardial ablation, incisions were closed, and endocardial ablation followed.

### Percutaneous Endocardial Ablation

Endocardial access to the left atrium was obtained via femoral vein access and conventional transseptal puncture guided by fluoroscopy or intracardiac echocardiography (ICE). Mapping and ablation catheters were introduced into the left atrium.

Since March 2013 NavX™ (Abbott, Abbott Park, IL, USA) was used to create electro anatomical LA maps to guide circumferential antral pulmonary vein isolation (PVI). Prior to March 2013 PVI was performed according to the ostial segmental isolation method.

While encircling the pulmonary vein ostia or antra the breakthroughs between epicardial lesions along the pericardial reflections were identified. Those breakthroughs were ablated with Cool Flex™ or Flexability™ (Abbott, Abbott Park, IL, USA) to complete the PVI. Sites with no voltage were identified as necrotic tissue from previous epicardial ablation and were therefore not ablated. Entrance block by circular mapping catheter confirmed PVI.

### Postoperative Management

If patients were not in sinus rhythm at the end of CP, electrical cardioversion was performed in the operating room. Other electrical or pharmacologic cardioversions and repeat catheter ablations were directed by the referring physician.

Initiation and discontinuation of antiarrhythmic drug therapy (AAD) and anticoagulation therapy throughout the follow up was managed by referring physician. Anticoagulation therapy was reinitiated after the CP for at least 3 months.

After February 2010 all the patients received esophagoscopy before discharge (18).

### Follow-Up Monitoring

The outcome was assessed at one year after the procedure and at long-term follow up by clinical presentation and rhythm monitoring.

#### 1-year follow up

61.2% (71/116) of included patients were implanted with an implantable loop recorder (ILR [Reveal™ XT Insertable Cardiac Monitors; Medtronic, Minneapolis, MN] or [Reveal LINQ™ Insertable Cardiac Monitors; Medtronic, Minneapolis, MN]) right after CP. The rhythm and daily AF burden (AFB = % of time [day] in AF) were stored in ILR and then downloaded and assessed at 1-year ambulatory visit. 38.8% (45/116) patients did not receive ILR. For these patients, rhythm at 1-year follow up was assessed by clinical presentation and with 12-lead ECG recording.

#### Long-term follow up

Long-term follow up was performed between October 2021 and December 2022. Patients’ cardiology documentation was examined; patients were classified depending on clinical presentation and 12-lead ECG recording at previous ambulatory visits. Patients who had sinus rhythm at previous ambulatory visits were monitored with 7-day ECG Holter. AF burden was determined. 8.4% (10/119) of patients having sinus rhythm on 12-lead ECG recording at last ambulatory visit, refused a 7-day Holter ECG. For these patients, rhythm at long-term follow up was assessed by clinical presentation and with 12-lead ECG recording. Patients with unclear history of AF were telephoned and examined on ambulatory visit. They were monitored with a 7-day Holter. AF burden was determined. Patients having clinical history of persistent AF on previous visits had telephone call for demographic characteristics but were not evaluated with a 7-day Holter. Class Ic and III AADs were assessed at 1-year follow up and at long-term follow up. Repeat catheter ablations prior to long-term follow up were documented.

### Definition of procedural endpoint and outcomes

Procedural endpoint for epicardial part of CP was visual overlapping of ablation lesions on posterior wall of the left atrium. Procedural success for epicardial part of CP was isolation of pulmonary veins confirmed with entrance block by circular mapping catheter.

One-year success for patients with persistent AF was defined as freedom from AF or AFL (atypical flutter) on 12-lead ECG recording and AF/AFL burden <1% on ILR. For paroxysmal AF any recorded and stored arrhythmia episode on ILR lasting at least 6 minutes was inspected and identified as recurrent AF or AFL.

Long-term success was defined as freedom from AF or AFL with sinus rhythm on 12-lead ECG recording and AF/AFL burden <1% on 7-day Holter ECG.

### Complications

Major periprocedural complications were defined as events related to the procedure that happened in less than 30 days and enlarged hospital stay, resulted in rehospitalization, intervention and/or had long-term negative impact on patients’ health. Minor periprocedural complications were defined as events related to the procedure that happened in less than 30 days but did not cause any of the conditions listed above. Major long-term adverse events were defined as events related to the procedure and happened in 30 days or later from the CP and required rehospitalization, intervention and/or had long-term negative impact on patients’ health.

For prevention of acute postprocedural pericarditis all patients received an intravenous bolus of steroids at the end of the CP and NSAIDs orally were prescribed for at least two weeks.

### Statistical Analysis

Normally distributed numeric measures are reported as mean ± standard deviation (SD) and non-normally distributed variables as median ± interquartile range. Categorical measures are presented as counts and percentages.

Kaplan-Meier survival analysis was calculated for AF recurrence.

Multivariate Cox Regression analysis with 95% confidence interval was performed to assess predictors of outcome. A p-value <0.05 was considered statistically significant. For statistical analysis SPSS version 24.0 (IBM Corporation, Armonk, NY) was used.

## Results

### Enrolment and Demographic Characteristics

170 patients underwent CP from January 2009 until July 2020. 47 were excluded; 38 due to lack of long-term follow up and 13 due to death before long-term follow up. In 11 patients death was not related to the ablation procedure. As reported previously 2 patients died from atrio-esophageal fistula in the first 20 cases; after that additional safety mechanisms were implemented in the procedure (18,19). Additionally, 4 patients with only epicardial part of CP were also excluded.

119 patients with endocardial and epicardial ablation and long-term-follow up were included in the analysis. 90.8% (108/119) of patients underwent CP in a single-setting and 9.2% (11/119) were treated in staged CP. The access to pericardial space was obtained through laparoscopic transabdominal transdiaphragmatic approach in 96.9% of patients (115/119) and through subxiphoid approach in 3.4% of patients (4/119).

Demographic characteristics are described in Table 1. Average age at CP was 58.4 ± 8.0 years and average age at long-term follow up was 66.7 ± 7.3 years. Majority of patients were male (77%). 70.6% (84) of patients had longstanding persistent AF, 5.9% (7) had persistent AF and 23.5% (28) paroxysmal AF, as defined by the recommendations of Heart Rhythm Society (20) and European Society of Cardiology (21). Average duration of AF before CP was 4.9 ± 4.5 years.

**Table 1.** Baseline Demographic Characteristics at CP.

| Demographic Characteristics | Baseline at CP |
| --- | --- |
| Male, N (%) | 92 (77%) |
| Age, years (SD) | 58.4 (8.0) |
| Body mass index, kg/m<sup>2</sup> (SD) | 29.6 (4.9) |
| AF duration, years (SD) | 4.9 (4.1) |
| AF type, N (%) |  |
| Persistent | 7 (5.9) |
| Longstanding persistent | 84 (70.6) |
| Paroxysmal | 28 (23.5) |
| Type of CP, N |  |
| CP-Single-Setting | 108 (90.8) |
| CP-Staged | 11 (9.2) |
| Chronic heart failure, N (%) | 17 (14.3) |
| Arterial Hypertension History, N (%) | 78 (65.5) |
| Vascular disease, N (%) | 8 (6.7) |
| Stroke/TIA/thromboembolic Event, N (%) | 2 (1.7) |
| Diabetes Mellitus, N (%) | 11 (9.2) |
| CHA2DS2VASc, N (%) |  |
| CHA2DS2VASC 0 | 25 (21.0) |
| CHA2DS2VASC 1 | 46 (38.7) |
| CHA2DS2VASC 2 | 30 (25.2) |
| CHA2DS2VASC 3 | 10 (8.4) |
| CHA2DS2VASC 4 | 7 (5.9) |
| CHA2DS2VASC 5 | 1 (0.8) |
| CHA2DS2VASC 6 | 0 (0.0) |
| CHA2DS2VASC 7 | 0 (0.0) |

At baseline 40.3% of patients had CHA2DS2VASc score ≥2; 14.3% had history of heart failure, 65.5% arterial hypertension, 6.7% vascular disease (prior myocardial infarction, coronary artery disease, peripheral artery disease), 1.7% prior stroke/TIA/thromboembolic event and 9.2% diabetes mellitus. At long-term follow up 72.3% of patients had CHA2DS2VASc score ≥2; 37% had history of heart failure, 76% arterial hypertension, 17% vascular disease, 2% prior stroke/TIA/thromboembolic event and 13% diabetes mellitus.

Median follow up one year after CP was 1.1 ± 0.3 years. Median long-term follow up was 8.3 ± 2.8 years; the longest follow up was 13.5 years after CP.

Demographic characteristics at long-term follow up are described in Table 2.

**Table 2.**
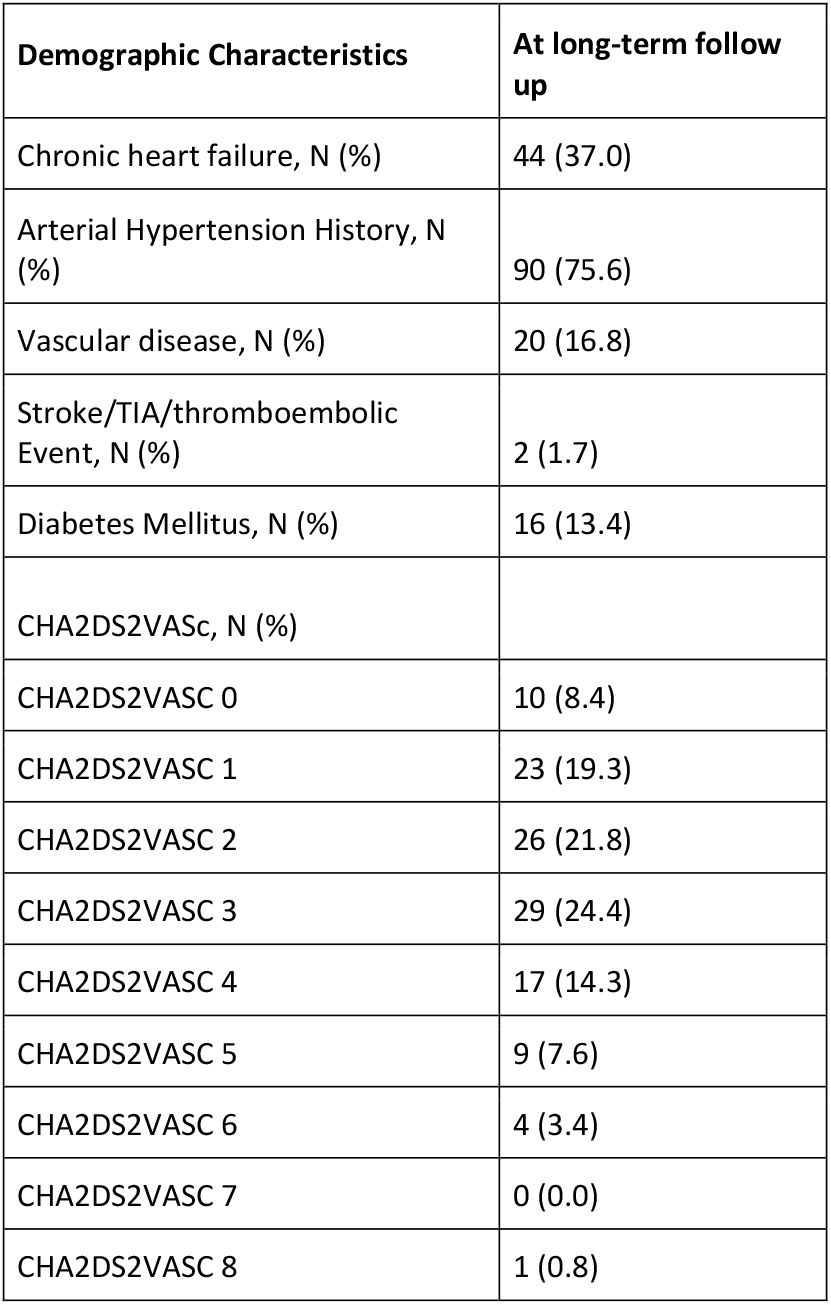
Demographic Characteristics at long-term follow up.

| **Demographic Characteristics** | **At long-term follow up** |
| --- | --- |
| Chronic heart failure, N (%) | 44 (37.0) |
| Arterial Hypertension History, N (%) | 90 (75.6) |
| Vascular disease, N (%) | 20 (16.8) |
| Stroke/TIA/thromboembolic Event, N (%) | 2 (1.7) |
| Diabetes Mellitus, N (%) | 16 (13.4) |
| CHA2DS2VASC, N (%) |  |
| CHA2DS2VASC 0 | 10 (8.4) |
| CHA2DS2VASC 1 | 23 (19.3) |
| CHA2DS2VASC 2 | 26 (21.8) |
| CHA2DS2VASC 3 | 29 (24.4) |
| CHA2DS2VASC 4 | 17 (14.3) |
| CHA2DS2VASC 5 | 9 (7.6) |
| CHA2DS2VASC 6 | 4 (3.4) |
| CHA2DS2VASC 7 | 0 (0.0) |
| CHA2DS2VASC 8 | 1 (0.8) |

## Outcomes

### 1-year follow up

1-year follow up data was available for 116 patients; 3 included patients did not attend 1-year follow up. 91.4% (106/116) of patients had sinus rhythm on 12-lead ECG and 8.6% (10/116) had AF or AFL. 73.6% (78/106) of patients in sinus rhythm were free off AADs.

61.2% (71/116) patients had implanted ILR. Of those patients who had implanted ILR, 76.1% (54/71) had AF or AFL burden <1% on ILR. 23.9% (17/71) had AF or AFL burden ≥1%. 79.6% (43/54) of patients with AF or AFL burden <1% were free off AADs.

Of all patients in the analysis 45.4% (54/119) had AF or AFL burden <1%, 14.3% (17/119) had AF or AFL burden ≥1%. For 40.3% (48/119) there was not available AF or AFL burden data.

No additional catheter ablations were performed in the first year.

### Long-term follow up

Long-term follow up data was available for all included patients (119). 65.5% (78/119) of patients had sinus rhythm on 12-lead ECG and 34.5% (41/119) had AF or AFL. 85.9% (67/78) of patients in sinus rhythm on 12-lead ECG were free off AADs 1c or III class. Additional catheter ablations were performed in 44.5% (53/119) of all patients, including additional catheter ablations in 34.6% (27/78) of patients who had sinus rhythm on 12-lead ECG.

Of all included patients, 53.8% (64/119) had AF or AFL burden <1% on 7-day Holter ECG. 87.5% (56/64) of patients with AF or AFL burden <1% on 7-day Holter ECG were free off AADs 1c or III class. Additional catheter ablations were performed in 32.8% (21/64) of patients who had AF or AFL burden <1% on 7-day Holter ECG.

Time-to-recurrence Kaplan-Mayer analysis for AF or AFL is shown in Figure 3.

**Figure 3:**
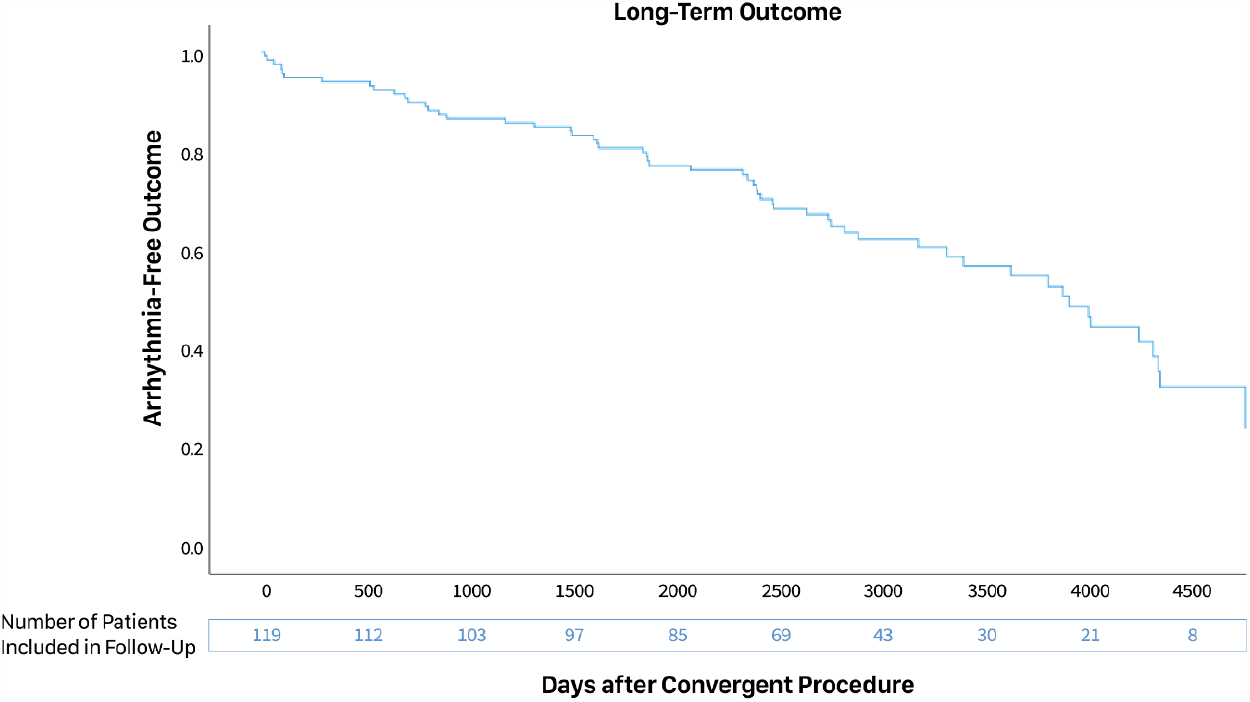
AF or AFL-Free Outcome in Patients after CP (time-to-recurrence Kaplan-Mayer analysis)

### Predictors of Recurrence

Multivariate Cox Regression analysis did not identify any predictors significantly associated with increased risk of AF recurrence after one year.

|  | Sig. | Exp(B) | 95.0% CI for Exp(B) |  |
| --- | --- | --- | --- | --- |
|  |  |  | Lower | Upper |
| Age | .634 | 1.037 | .892 | 1.207 |
| Gender | .678 | 1.815 | .109 | 30.273 |
| BMI (kg/m2) | .276 | 1.135 | .904 | 1.426 |
| LAVI (ml/m2) | .222 | 1.070 | .960 | 1.192 |
| LVEF (%) | .127 | .944 | .877 | 1.016 |
| PAF | .260 | .226 | .017 | 3.004 |
| PeAF | .991 | 404858.292 | .000 | . |
| LSPeAF | . |  |  |  |
| AF duration (years) | .556 | .913 | .675 | 1.235 |
| CHF | .588 | 3.285 | .044 | 243.742 |
| Hypertension History<br>(1) | .740 | 1.768 | .061 | 51.356 |
| Diabetes Mellitus<br>History<br>(1) | .711 | .612 | .045 | 8.231 |
| Stroke / TIA / TE<br>(2) | .608 | 2.864 | .051 | 160.226 |
| Vascular disease CAD,<br>PAD<br>(1) | . |  |  |  |
| CHA2DS2VASC score<br>(Max -10) | .294 | 2.942 | .392 | 22.073 |

At long-term follow up multivariate Cox Regression analysis identified three predictors associated with increased risk of AF recurrence: age (HR 1.15, 95% CI 1.02–1.29, P = 0.023), BMI (HR 1.31, 95% CI 1.05–1.63, P = 0.018) and LAVI (HR 1.08, 95% CI 1.01–1.15, P = 0.030). Other predictors were not significantly associated to AF recurrence.

|  | Sig. | Exp(B) | 95.0% CI for Exp(B) |  |
| --- | --- | --- | --- | --- |
|  |  |  | Lower | Upper |
| Age | .023 | 1.148 | 1.019 | 1.294 |
| Gender | .981 | .965 | .049 | 18.862 |
| BMI (kg/m2) | .018 | 1.308 | 1.048 | 1.633 |
| LAVI (ml/m2) | .030 | 1.078 | 1.007 | 1.154 |
| LVEF (%) | .511 | .979 | .921 | 1.042 |
| PAF | .669 | 1.565 | .200 | 12.232 |
| PeAF | .149 | .068 | .002 | 2.618 |
| LSPeAF | . |  |  |  |
| AF duration (years) | .312 | .892 | .714 | 1.114 |
| CHF | .271 | .141 | .004 | 4.619 |
| Hypertension History<br>(1) | .343 | 4.363 | .207 | 91.878 |
| Diabetes Mellitus History<br>(1) | .655 | 2.995 | .024 | 366.244 |
| Stroke / TIA / TE<br>(2) | .934 | 1.182 | .023 | 61.633 |
| Vascular disease CAD,<br>PAD<br>(1) | .238 | 18.499 | .145 | 2357.063 |
| CHA2DS2VASC score<br>(Max -10) | .573 | 1.743 | .252 | 12.063 |

### Complications

A total periprocedural major complication rate of 10.9% was observed. Three patients developed cardiac tamponade (2.5%), one with cardiogenic shock intraoperatively. One patient developed ventricular fibrillation intraoperatively that required CPR and electrocardioversion (0.84%). Three patients had hemorrhage (2.5%); one from abdominal wall that resulted in exploratory laparotomy and blood transfusion with 4 units, one was bleeding intraabdominally due to abdominal adhesions from a previous cholecystectomy, which required surgical evacuation of hematoma and transfusion of 2 units of blood and one hematemesis related to previously unknown gastric ulcer. One patient with bilateral pneumonia required rehospitalization (0.84%). One patient with pseudoaneurysm of superficial femoral artery was treated surgically (0.84%). Right phrenic nerve paresis was observed in one patient (0.84%) and ulnar nerve compression (0.84%) in one patient. One patient developed septic shock one day after procedure, which resolved with appropriate therapy (0.84%). One patient had acute decompensation of chronic cor pulmonale that resolved with therapy (0.84%). On esophagoscopy three patients had possible damage to the anterior wall of esophagus that resolved into localized small scar in couple of days after therapy with PPI and parenteral feeding. No patients experienced stroke during follow up.

Ten patients developed minor periprocedural complications (8.4%). One developed non-severe pulmonary embolism from previously unknown preoperative deep vein thrombosis (0.84%). Nine patients had ventral hernia (7.6%). One patient had hiatal hernia (0.84%). Two patients had small scar in esophagus and two small hematoma, that did not require any additional actions. In three patients gastritis was found. Due to prevention therapy of acute postprocedural pericarditis, this complication is not reported.

Major long-term adverse events rate was 2.5% as three ventral hernias required surgery later.

## Discussion

Our retrospective study showed that CP in our mixed group of patients with AF resulted in high probability of sinus rhythm maintenance of 76.1% at one year and 53.8% at long-term follow up after 8.3 years. Majority of patients in sinus rhythm (79.6% at one year and 87.5% at long-term follow up) were off AADs. We found that age, BMI and LAVI were predictors of long-term outcome. AF recurrence importantly increased with time. To maintain sinus rhythm at long-term follow up 32.8% of patients required at least one additional catheter ablation.

The multicenter, randomized Convergent trial demonstrated superiority of CP in patients with longstanding persistent AF with 73.7% effectiveness regardless of AAD use compared to 44.4% in the catheter ablation arm at 12 months (22). Recently published randomized clinical trial HARTCAP-AF showed 89% and 36% freedom from any supraventricular tachyarrhythmia lasting >30 seconds off AADs at 12 months after convergent ablation and catheter ablation respectively (23). Similarly, Varzaly et al. performed a meta-analysis where sinus rhythm maintenance at 79.4% was demonstrated after CP at 19 months follow up (24).

Importantly, outcomes for sinus rhythm maintenance after CP have been shown to decline over years. In a recent meta-analysis Eranki et al. showed freedom from AF of 78.2% and 73.6% at 1 and 3 years after CP respectively (16). In the latest study Pannone et al. reported ATs free survival without AADs in non-paroxysmal AF 76.7% at 12 months, 67.5% at 24 months, 60.5% at 36 months, 53.6% at 48 months, and 46.1% at 60 months (17).

Our results for short-term and mid-term outcome are consistent with studies mentioned above and previous studies from our group (14,18,25). However, our current study reports the longest follow-up after CP, with 53.8% freedom from AF/AFL recurrence at a median of 8.3 years.

Long-term results that are available for catheter ablation demonstrate low success rate in catheter ablation of persistent and long-term persistent AF. In Hamburg study, success rate of 45% was reported after multiple ablation procedures at ≥50 months (26). In a meta-analysis Ganesan et al. reported 3-year freedom from AF in patients with non-paroxysmal AF at 41.6% and 77.8% following a single catheter ablation and multiple procedures respectively (27). However, most of the studies included in meta-analysis performed follow up with 24-hour Holter, transtelephonic ECGs and some only performed symptom-related follow up, which is a less accurate method of detecting AF recurrence. The analysis of CIRCA-DOSE trial showed that sensitivity of detecting AF recurrence increased with the duration of rhythm monitoring. Serial short duration holter ECG monitoring (24-/48-hour) missed a substantial number of recurrences and less accurately estimated AF burden, however serial long-term monitors (14-day) had sensitivity closer to gold standard of continuous ECG monitoring (28). It is assumed that non-pulmonary vein substrate, located mainly on posterior wall of the left atrium and left atrial remodeling significantly contribute to sustaining arrhythmia in persistent and long-standing persistent AF (29,30). More extensive and durable transmural lesions of posterior wall with direct ablation of epicardial connections between endo- and epicardium and autonomic ganglia located in epicardial fat pads (31) resulting in homogenization of posterior wall (32) could play a major role in maintaining a sinus rhythm after CP.

### Complications

We report 10.9% of periprocedural major complication rate, 8.4% of minor periprocedural complications and 2.5% of major long-term adverse events. The complication rate is high, however our cohort included patients from the beginning of implementation of the CPs in our center. As previously reported in first 20 patients who underwent CP two patients developed atrio-esophageal fistulas and died. An extensive hospital investigation resulted in implementation of additional safety measures such as esophageal temperature monitoring, termination of RF energy dictated by esophageal temperature increase, cooling of pericardial space during epicardial ablation, minimization of endocardial ablation along the posterior wall of left atrium, implementation of esophagoscopy (19). We believe that these modifications by our group, implemented since February 2010, resulted in widespread use of CP on a global level, because they clearly demonstrated safety from atrio-esophageal fistulas, if the CP was done in a single setting. Since then, no atrio-esophageal fistulas or deaths occurred. Our results are consistent with previous research, which show major complication rate after CP between 0 and 20.8%, with a pooled complication rate of 5.53% (16,33–35). In a single center study Larson et al. showed that transitioning from transdiaphragmatic to subxiphoid approach enhances safety profile and significantly reduces complications. They reported major complication rate of 23% and 3.8% after CP with transdiaphragmatic or subxiphoid approach, respectively. Most complications were related to entering the abdominal cavity (31). However, subxiphoid approach results in poorer accessibility to the anterior segments of pulmonary veins hence only posterior wall of left atrial can be ablated epicardialy, which could reduce procedure’ success. Selecting patients for the CP, the expected risks of the procedure and benefits of sinus rhythm maintenance must be thoughtfully weighed.

### Limitations

Our study has some obvious limitations. Firstly, it is a single-center retrospective study, which somewhat reduces the importance of our conclusions. Secondly, there was some possible bias related to the inclusion of patients as a substantial number of patients that had received CP in our center refused to perform long-term ambulatory visit. Thirdly, although our study included a relatively large sample size, data for calculations of predictors of recurrence were available only for a smaller sample. Fourthly, for simplification purposes a cut off of <1% AF burden was used as absence of AF in analysis of ILR and 7-day holter recordings. The used cut off might correlate better with clinical outcomes, however, if a stricter cut off of any 30 s AF episode was used instead, it might increase the detection of AF recurrences. On the other hand, our group clearly demonstrate the evolution of CP from its initial phase to mature procedure, which was actually implemented in major centers around the world.

## Conclusions

CP resulted in high long-term probability of sinus rhythm maintenance. During long-term follow-up additional catheter ablations were required to maintain sinus rhythm in a substantial number of patients.

## Data Availability

Data will be available from authors on reasonable request.

